# Ocular diagnostic patterns among patients attending community outreach eye camps in Bangladesh: a retrospective registry-based study

**DOI:** 10.64898/2026.08.15.26360489

**Authors:** Md. Saleh Ahmed, Moutushi Islam, Anthony Albert, Akhter Ferdoshe Jahan, Nusrat Lubna Islam, Tasruba Shahnaz, Chowdhury Mashrur Mahdee

**Author notes:** Corresponding Author: Chowdhury Mashrur Mahdee, Research and Academics Department, Bashundhara Eye Hospital and Research Institute; Mailing address: Plot #474, Block-D, Sabrina Sohban Sharak, Bashundhara R/A, Dhaka-1229, Bangladesh.

## Abstract

**Introduction:** Community outreach eye camps extend eye care to underserved rural populations across South Asia, but published data mostly describe surgical yield rather than the full range of presenting conditions. This study described the pattern of ocular diagnoses among patients attending community eye camps in Bangladesh and examined how these patterns varied by year, age, and sex.

**Methodology:** This was a retrospective, registry-based study of patients screened through the outreach eye-camp programme of Bashundhara Eye Hospital and Research Institute, Bangladesh, between 2020 and 2024 (2021 excluded due to COVID-19). Age, sex, and provisional diagnosis were recorded for every patient and grouped into five categories: cataract and lens disorders, refractive errors and presbyopia, ocular surface inflammatory disorders, lacrimal system disorders, and miscellaneous disorders. Multivariable logistic regression models estimated the adjusted odds and predicted prevalence of each diagnostic category by calendar year (adjusted for age group and sex) and by age group and sex (adjusted for calendar year).

**Results:** Among 7,267 patients, refractive errors and presbyopia were the most common category (51.31%), followed by cataract and lens disorders (23.54%), miscellaneous disorders (13.86%), ocular surface inflammatory disorders (8.59%), and lacrimal disorders (2.70%). Ocular surface disorders declined significantly over time (adjusted OR: 0.80/year; p-value<0.001). Cataract showed a borderline decline (adjusted OR: 0.94; p-value=0.065). Cataract prevalence rose steeply with age, from 3-6% below 18 years to 49-55% at 60 and older, while refractive errors/presbyopia peaked at 40-59 years (64-68%) before declining thereafter (30-37%). Ocular surface disorders were most frequent among children. Lacrimal disorders remained uncommon (2-5%), with no significant interaction.

**Conclusion:** Refractive errors and presbyopia were the leading reason for presentation, with distinct age patterns across diagnostic categories. Outreach eye camps should be resourced for both comprehensive refraction, primary eye care and cataract-surgical referral, reflecting actual community-level diagnostic needs.

## Introduction

Vision impairment remains one of the most widespread and under-addressed public health problems worldwide. According to the World Health Organization at least 2.2 billion people globally live with some form of vision impairment, and in at least 1 billion of these cases the impairment could have been prevented or has yet to be addressed [1]. Uncorrected refractive error is the leading cause of vision impairment worldwide, ahead of cataract, age-related macular degeneration, and glaucoma [1,2]. Non-blinding disorders such as dry eye and conjunctivitis are also among the most frequent reasons patients seek eye care in every country, and are increasingly recognized as an important part of the overall eye-care burden [1]. Presbyopia alone is estimated to affect around one in four people globally, with many left with uncorrected near vision impairment because they lack access to reading glasses [3]. This evidence indicates that the population-level demand for eye care extends well beyond cataract and other blinding conditions, and includes a large number of refractive, presbyopic, and ocular-surface complaints that are comparatively simple and inexpensive to manage.

Bangladesh follows this broader global pattern, though historically its eye-care planning has centered heavily on cataract because of its contribution to blindness. The National Blindness and Low Vision Survey of Bangladesh found that cataract accounted for approximately 80% of bilateral blindness among adults 30 years and older, most of it treatable with timely surgery [4]. A more recent national cross-sectional survey confirmed that cataract remains the single largest contributor to blindness and low vision in Bangladeshi adults, with a substantial population-attributable risk even after accounting for other causes such as age-related macular degeneration and diabetic retinopathy [5]. However, community-level studies suggest that the true burden of eye disease in Bangladesh is broader than cataract alone. A survey of an urban slum population in Dhaka found that refractive error and cataract together accounted for the majority of eye disease, and highlighted that eye conditions in general remain a neglected health issue with limited public awareness [6]. Similarly, a rural district-based study found generally poor community awareness of common eye conditions beyond cataract, pointing towards a wider unmet need for screening and education across the range of ocular disorders [7].

Community outreach eye camps have long served as a practical model for extending eye-care access to rural and underserved populations across South Asia, where formal eye-care infrastructure is concentrated in cities and surgical backlog is high. Camp-based screening followed by centralized, low or no cost surgery has been described in Indian eye camps [8], mass cataract-surgery campaigns in other developing-country settings [9], and district-level needs assessments in Bangladesh [10,11]. Displaced and host-community populations have also been reached through this model, as shown in studies conducted among Rohingya refugees and their host communities in Cox’s Bazar [12,13]. In all of these studies, the primary outcome of interest has almost always been cataract-surgical coverage or postoperative visual outcome. Far less attention has been paid to describing the full diagnostic case-mix of patients who present at these camps, many of whom are screened, examined, and managed for conditions other than cataract, such as refractive error, presbyopia, and ocular surface disease.

To the best of our knowledge, no prior study has systematically described the distribution of ocular diagnoses among patients attending community outreach eye camps in Bangladesh, or examined how this distribution varies with calendar year, age, and sex.

The purpose of this study was to describe the pattern of ocular diagnoses among patients attending the community outreach eye-camp programme, and to evaluate how the prevalence of major diagnostic categories varied by calendar year, age group, and sex, in order to inform more evidence-based planning of eye-camp staffing, equipment, and service delivery.

## Methodology

### Study design, setting, and data collection

This was a retrospective, registry-based study of patients screened through the outreach eye-camp programme of Bashundhara Eye Hospital and Research Institute (BEHRI), a tertiary eye care facility based in Dhaka, Bangladesh. As part of its community outreach activities, BEHRI regularly conducts free eye camps at rural sites across multiple districts of Bangladesh, with several camps typically held in each district. Camps were organized following prior discussions with local organizers and community partners to determine suitable sites and dates. Around two weeks before each camp, community-level promotional activities and patient registration began. Promotional activities included miking (announcements via loudspeaker), poster displays, and announcements at local mosques to inform residents of the upcoming free eye camp. Interested individuals registered in advance and were screened at the camp.

At each camp, a consultant ophthalmologist and a team of nurses and ophthalmic assistants travelled from BEHRI with portable diagnostic equipment to conduct on-site screening. Basic demographic data (age and sex) and a provisional ocular diagnosis were recorded for every patient screened at the camp. This camp-level screening register is the data source used for the present study. This model of camp-based community screening is widely used to deliver eye care and expand coverage in under-served and rural populations of South Asia [8,9], including previous outreach-based eye care delivery models conducted in Bangladesh [10–13].

All camp-screening data were entered and stored securely in BEHRI’s institutional database. For the purposes of this study, screening records from camps conducted between 2020 and 2024 were retrospectively extracted, 2021 was excluded as no camps were conducted that year because of the COVID-19 pandemic. After excluding records with incomplete information, 7,267 patients with complete information were included in the final sample. This study was conducted in accordance with the tenets of the Declaration of Helsinki. Informed consent was obtained from all patients. The study received an exemption from full ethical review by the Institutional Review Board of Bashundhara Eye Hospital and Research Institute (BEHRI-IRB/2026/0803), Dhaka, Bangladesh.

### Diagnostic classification

Provisional diagnoses were recorded at the camp as free-text clinical entries, which yielded a large number of individual diagnostic labels. To make these entries usable for analysis, all original diagnostic labels were reviewed and grouped, based on shared anatomical site and clinical similarity, into five mutually exclusive diagnostic categories: cataract and lens disorders, refractive errors and presbyopia, ocular surface inflammatory disorders, lacrimal system disorders, and miscellaneous ocular disorders, a residual category comprising all remaining, individually low-frequency diagnoses. The full mapping of original registry entries to these five categories is provided in the supplementary table.

### Statistical analysis

Descriptive statistics were used to summarize the demographic characteristics of the study population and the overall distribution of diagnostic categories. For each of the four specific diagnostic categories other than the miscellaneous group, a multivariable logistic regression model was fitted with the presence of that diagnostic category (yes/no) as the outcome, calendar year modelled as a continuous variable as the primary exposure, and age group and sex included as covariates. The adjusted odds ratio (OR) for each additional calendar year, with 95% confidence interval (CI) and p-value, was obtained from each model. Adjusted predicted prevalence of each diagnostic category for each calendar year was derived from the same models and compared descriptively with the observed (unadjusted) prevalence for that year. As the miscellaneous ocular disorders category was a non-specific combination of low-frequency diagnoses instead of a clinically coherent entity, it was only analyzed descriptively and not modelled.

To evaluate whether diagnostic-category prevalence varied jointly by age and sex, a second set of multivariable logistic regression models was fitted for each of the four specific diagnostic categories, replacing the continuous year term with an age group × sex interaction term, adjusting for calendar year. Adjusted predicted prevalence for each age group × sex stratum was obtained from these models, and the statistical significance of the age group × sex interaction was assessed for each diagnostic category. A p-value<0.05 was considered statistically significant throughout the analysis. All analyses were conducted using Stata (version 17.0; StataCorp LP, College Station, Texas).

## Results

Between 2020 and 2024 (excluding 2021), 7,267 patients with complete demographic and diagnostic information were screened through BEHRI’s outreach eye-camp programme and included in this analysis. Table 1 shows the demographic characteristics of the study population. Screening activity was unevenly distributed across years, with 2023 alone accounting for nearly two-thirds of all records (4,704; 64.73%), followed by 2024 (1,344; 18.49%), 2022 (747; 10.28%), and 2020 (472; 6.50%). Majority of patients screened were female (4,178; 57.49%). By age, 45.03% of patients were between 40-59 years, 31.33% were 60 years or older, 18.15% were between 18-39 years, and 5.49% were younger than 18 years.

**Table 1:** Baseline demographic characteristics of patients attending community eye camps between 2020 and 2024 (N = 7,267).

| Characteristics | n (%) |
| --- | --- |
| **Year** |  |
| 2020 | 472 (6.50) |
| 2022 | 747 (10.28) |
| 2023 | 4704 (64.73) |
| 2024 | 1344 (18.49) |
| **Sex** |  |
| Male | 3089 (42.51) |
| Female | 4178 (57.49) |
| **Age group** |  |
| <18 | 399 (5.49) |
| 18-39 | 1319 (18.15) |
| 40-59 | 3272 (45.03) |
| 60+ | 2277 (31.33) |
| **Total** | **7267** |

Table 2 shows the overall distribution of ocular diagnostic categories among the 7,267 patients screened. Refractive errors and presbyopia were the largest category, accounting for just over half of all patients (3,729; 51.31%). Cataract and lens disorders were the second most common category (1,711; 23.54%), followed by miscellaneous ocular disorders (1,007; 13.86%), ocular surface inflammatory disorders (624; 8.59%), and lacrimal system disorders (196; 2.70%).

**Table 2:** Distribution of diagnostic categories among patients attending community eye camps (N = 7,267).

| Diagnostic category | n (%) |
| --- | --- |
| Cataract and lens disorders | 1711 (23.54) |
| Refractive errors and presbyopia | 3729 (51.31) |
| Ocular surface inflammatory disorders | 624 (8.59) |
| Lacrimal system disorders | 196 (2.70) |
| Miscellaneous ocular disorders | 1007 (13.86) |
| **Total** | **7267** |

Table 3 shows the adjusted temporal trend for each diagnostic category, expressed as the adjusted OR per calendar year from logistic regression models adjusted for age group and sex. Ocular surface inflammatory disorders showed a statistically significant decline over the study period (adjusted OR: 0.80; 95% CI: 0.74, 0.87; p-value<0.001). Cataract and lens disorders (adjusted OR: 0.94; 95% CI: 0.88, 1.00; p-value=0.065), refractive errors and presbyopia (adjusted OR: 1.04; 95% CI: 0.98, 1.09; p-value=0.168), and lacrimal system disorders showed no significant trend (adjusted OR: 0.94; 95% CI: 0.81, 1.09; p-value=0.423).

**Table 3:** Adjusted temporal trends in ophthalmic diagnostic categories estimated using logistic regression with calendar year modeled as a continuous variable.

| Diagnostic category | OR (95% CI) per year | p-value |
| --- | --- | --- |
| Cataract and lens disorders | 0.94 (0.88, 1.00) | 0.065 |
| Refractive errors and presbyopia | 1.04 (0.98, 1.09) | 0.168 |
| Ocular surface inflammatory disorders | 0.80 (0.74, 0.87) | <0.001 |
| Lacrimal system disorders | 0.94 (0.81, 1.09) | 0.423 |

Odds ratios represent the average annual change in the odds of each diagnosis. Models were adjusted for age group and sex.

Table 4 shows the observed and adjusted predicted prevalence of each diagnostic category by calendar year. The predicted prevalence of cataract and lens disorders fell from 28.51% in 2020 to 22.58% in 2022 and remained broadly stable ever since (23.10% in 2023, 23.97% in 2024). Refractive errors and presbyopia rose sharply from 42.13% in 2020 to a peak of 60.61% in 2022, before declining to around half of all patients in the two following years (50.94% in 2023, 50.69% in 2024). Ocular surface inflammatory disorders fell significantly, from 16.43% in 2020 to 6.98% in 2022, and remained in a relatively low in 2023 (8.31%) and 2024 (7.46%). Lacrimal system disorders remained low across the four years, fluctuating between 1.32% and 3.49%.

**Table 4:** Observed and adjusted predicted prevalence (%) of ophthalmic diagnostic categories by calendar year from multivariable logistic regression models.

| Diagnostic category | Estimate | Year |  |  |  |
| --- | --- | --- | --- | --- | --- |
|  |  | 2020 (%) | 2022 (%) | 2023 (%) | 2024 (%) |
| Cataract and lens disorders | Observed | 27.75 | 23.69 | 22.92 | 24.18 |
|  | Adjusted | 28.51*** | 22.58*** | 23.10*** | 23.97*** |
| Refractive errors and presbyopia | Observed | 40.89 | 59.97 | 51.02 | 51.19 |
|  | Adjusted | 42.13*** | 60.61*** | 50.94*** | 50.69*** |
| Ocular surface inflammatory disorders | Observed | 17.80 | 6.69 | 8.35 | 7.22 |
|  | Adjusted | 16.43*** | 6.98*** | 8.31*** | 7.46*** |
| Lacrimal system disorders | Observed | 3.60 | 1.34 | 3.02 | 2.01 |
|  | Adjusted | 3.49*** | 1.32** | 3.03*** | 2.01*** |

\* = p-value < 0.05; \*\* = p-value < 0.005; \*\*\* = p-value < 0.001. Adjusted predicted probabilities were estimated from logistic regression models adjusted for age group and sex.

Table 5 and Figure 1 show the adjusted predicted prevalence of each diagnostic category by age group and sex, together with the statistical significance of the age group × sex interaction for each category. Cataract and lens disorders rose steeply with age in both sexes, from 3.17% (male) and 5.71% (female) among patients younger than 18 years, through 5.39% (male) and 2.46% (female) in the 18-39 year group and 13.61% (male) and 14.56% (female) in the 40-59 year group, to 55.22% (male) and 48.79% (female) among patients 60 years and older. This age × sex interaction was statistically significant (p-value<0.001). Refractive errors and presbyopia followed a different pattern, rising from around 42-52% in patients younger than 40 years to a peak in the 40-59 year age group (67.60% male, 64.28% female), before falling to 29.53% (male) and 36.76% (female) among patients 60 years and older. Age × sex interaction for refractive errors and presbyopia was also statistically significant (p-value<0.001). Ocular surface inflammatory disorders were most frequent among the youngest patients (29.10% male, 21.43% female in the under 18 group) and declining progressively with age to their lowest levels among patients 60 years and older (3.84% male, 3.79% female). Age × sex interaction for ocular surface inflammatory disorders was significant (p-value<0.001). Lacrimal system disorders remained low and relatively stable across all age-sex strata, ranging from 1.90% to 4.76%. However, the age × sex interaction for this category did not reach statistical significance (p-value=0.334).

**Figure 1:**
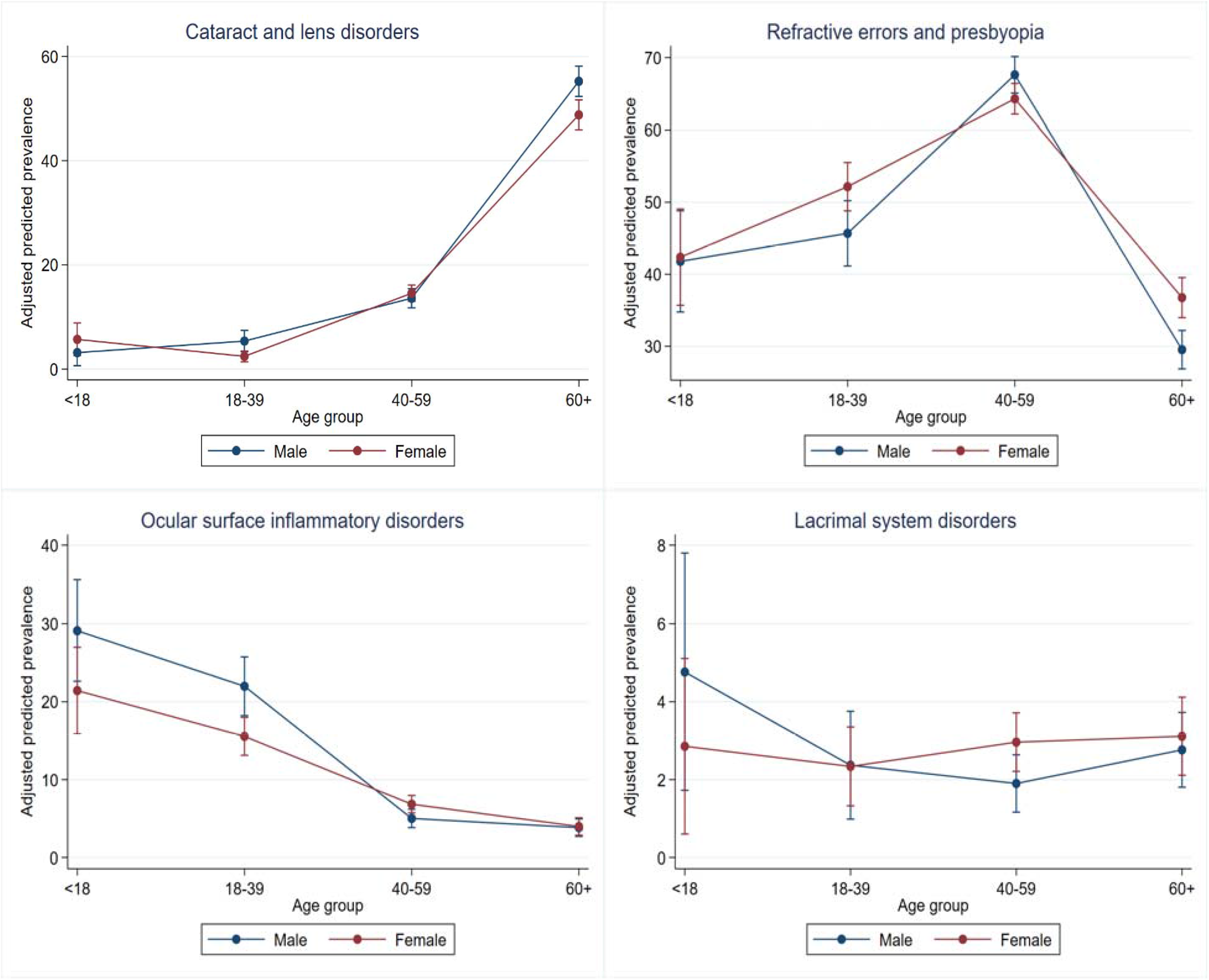
Adjusted predicted prevalence of diagnostic categories by age group and sex.

**Table 5:** Predicted prevalence (%) of diagnostic categories by age group and sex, with tests for age × sex interaction.

| Diagnostic category | Adjusted predicted prevalence (%) |  |  |  |  |  |  |  | Age × Sex interaction p-value |
| --- | --- | --- | --- | --- | --- | --- | --- | --- | --- |
|  | <18 M | <18 F | 18-39 M | 18-39 F | 40-59 M | 40-59 F | ≥60 M | ≥60 F |  |
| Cataract and lens disorders | 3.17 | 5.71 | 5.39 | 2.46 | 13.61 | 14.56 | 55.22 | 48.79 | <0.001 |
| Refractive errors and presbyopia | 41.80 | 42.38 | 45.69 | 52.16 | 67.60 | 64.28 | 29.53 | 36.76 | <0.001 |
| Ocular surface inflammatory disorders | 29.10 | 21.43 | 21.98 | 15.56 | 5.02 | 6.85 | 3.84 | 3.79 | <0.001 |
| Lacrimal system disorders | 4.76 | 2.86 | 2.37 | 2.39 | 1.90 | 2.96 | 2.77 | 3.11 | 0.334 |

## Discussion

This registry-based study of 7,267 patients attending community outreach eye camps in Bangladesh found that refractive errors and presbyopia were the leading reason for presentation, accounting for just over half of all diagnoses, which is more than double the proportion attributable to cataract and lens disorders. This finding is consistent with the broader global evidence base, in which uncorrected refractive error is recognized as the leading cause of vision impairment worldwide, and presbyopia alone is estimated to affect roughly a quarter of the global population [1–3]. Community and population-based studies from other Asian countries follow similar patterns. The Andhra Pradesh Eye Disease Study and the Singapore Epidemiology of Eye Disease Study have both identified refractive error as one of the most prevalent ocular conditions in general adult populations, occurring far more frequently than cataract when the entire population is examined [14,15]. Our results extend this observation specifically to the outreach camp setting in Bangladesh, and suggest that camps historically organized around cataract-surgical delivery are, in practice, seeing a caseload in which refraction and presbyopic correction represent the dominant day-to-day clinical need.

Cataract remains globally recognized as the most common treatable cause of vision loss [16]. Cataract and lens disorders represented a substantial share of the diagnostic case-mix in this registry (23.54%), heavily concentrated among older patients and rising from under 6% among those younger than 40 years to over half of patients aged 60 years and older. This age difference mirrors the well-established epidemiology of age-related cataract, where increasing age is consistently the single strongest risk factor for cataract [17,18]. It is also consistent with Bangladesh’s own national data, which have identified cataract as the single largest contributor to blindness and visual impairment in the country [4,5], and which originally motivated the emphasis on cataract-surgical delivery in outreach camp models such as the one described here and in comparable programmes in other South Asian eye camps [8,9] and in district-level assessments in Bangladesh [10,11]. The adjusted predicted prevalence of cataract and lens disorders declined over the study period, from 28.51% in 2020 to a more stable 23-24% from 2022 onward. A plausible explanation is that as the same outreach programme has performed a large volume of cataract surgery through its surgical referral pathway, the local pool of previously undiagnosed, surgically eligible cataract may have been progressively reduced, allowing camps to see relatively more of the underlying refractive and other ocular demand that would otherwise have been overshadowed. This remains speculative given the retrospective, non-experimental design of the current study, and would need to be tested in further studies.

Ocular surface inflammatory disorders showed a different age pattern from cataract, being most frequent among children and adolescents (21-29% among patients younger than 18 years) and declining steadily thereafter to under 4% among those 60 years and older. This pattern is broadly consistent with a community eye-health survey among school children in a rural Nigerian community, which found allergic conjunctivitis to be the single most common ocular disorder [19]. In contrast to the surgical referral pathway necessary for cataract in older patients, the concentration of ocular surface diseases among younger individuals has practical consequences for camp screening because most of these disorders can be treated medically on-site without further referral. We also observed a substantial decline in the overall prevalence over the study period, from over 16% in 2020 to below 9% from 2022 onward. Since environmental exposure and hygiene-related transmission have a significant impact on ocular surface inflammatory disease, this may partially account for the distinctive conditions of the 2020 camps, which were held in the early stages of the COVID-19 pandemic. Changes in hygiene practices and care-seeking patterns may have affected the actual prevalence of these conditions and the patients who arrived at camps. However, this explanation cannot be confirmed from the available registry data and requires further studies to be conclusive.

The distinct age pattern seen for refractive errors and presbyopia is unlikely to reflect a true decline in refractive need with age. A more probable explanation is that reliable refraction becomes harder to obtain and to classify once an eye has visually significant cataract. Population-based refractive error surveys routinely handle this by excluding such eyes altogether. A major US population-based study of refractive error excluded all eyes with cataract or prior cataract surgery before estimating prevalence, because refraction could not be meaningfully assessed through an opacified lens [20]. Camp-based screening is likely to face the same difficulty. As lens opacity increases with age, it becomes the more obvious and actionable finding, while the refractive component of a patient’s vision loss becomes harder to isolate and record. The decline in refractive errors and presbyopia diagnosis among the oldest patients is most likely due to this increasing difficulty of assessing refraction through a cataractous lens, instead of having fewer older patients requiring treatment.

Women made up a majority of patients screened in this registry (57.49%). Globally, the burden of eye conditions and vision impairment is not borne equally, and tends to fall disproportionately on women, older people, and rural or disadvantaged communities [1]. Community outreach screening has specifically been identified as a means of improving equity of access to eye care for women, alongside other marginalized groups [21], which may partly explain why women were the majority among patients attending the camps. Within most diagnostic categories the age pattern was broadly similar for men and women, with the age group × sex interaction reaching statistical significance for cataract and lens disorders, refractive errors and presbyopia, and ocular surface inflammatory disorders. This indicates that while the overall age gradient for each of these conditions differed somewhat in magnitude between the sexes, the general direction of the age pattern was shared.

Lacrimal system disorders were consistently the least common diagnostic category in this registry (2.70%) and were the only category for which the age group × sex interaction was not statistically significant. Lacrimal conditions in typical clinical populations include a congenital, pediatric component and a separate, acquired, older-adult component, which can produce a bimodal age distribution. The broad four-category age grouping used in this registry may not have been comprehensive enough to detect such pattern, which may partly explain the non-significance observed.

This study has several limitations. First, diagnoses were recorded as part of routine camp-based clinical screening rather than through a standardized research protocol. Second, the original free-text diagnostic entries were grouped into five categories post hoc by the study team, based on clinical and anatomical similarity, which is provided in full in the supplementary table. Some grouping decisions inevitably involve judgement, and a different grouping scheme could yield somewhat different category-level estimates. Third, screening volume was highly uneven across the study years, with 2023 contributing nearly two-thirds of all records. This imbalance limits the precision of estimates for the lower-volume years and means that year-specific findings should be interpreted cautiously. Fourth, patients attending free community eye camps are a self-selected population who chose to seek care, and camp location, promotional reach, and local demographic composition can all influence who presents. As a result, these findings describe the diagnostic pattern of care-seeking demand at this specific outreach programme, and should not be interpreted as a population-based estimate of eye disease prevalence in Bangladesh.

Despite these limitations, this study provides, to our knowledge, the first detailed description of the diagnostic case-mix among patients attending community outreach eye camps in Bangladesh. By characterizing the full range of conditions presenting at these camps, this study offers locally generated evidence to guide staffing, equipment, and decision-making for future outreach eye-care delivery in Bangladesh and similar settings.

## Conclusion

In this large, registry-based study of community outreach eye camps in Bangladesh, refractive errors and presbyopia were the leading reason for patient presentation. Cataract and lens disorders were concentrated among older patients and declined modestly over the study period. While ocular surface inflammatory disorders were concentrated among children, and lacrimal system disorders remained uncommon across all age and sex groups. These findings suggest that community eye camps should be resourced for comprehensive, age-appropriate primary eye care, including routine refraction and spectacle dispensing and on-site management of ocular surface disease in younger patients. Continued registry-based monitoring of diagnostic case-mix, alongside surgical outcome data, would help align camp staffing and equipment with actual community need over time.

## Data Availability

All data produced in the present study are available upon reasonable request to the authors

## Data availability statement

The data are available from the corresponding author upon reasonable request.

## Conflicts of interest

The authors declare that there is no conflict of interest regarding the publication of this article.

## Funding statement

This research did not receive any specific grant from funding agencies. The study was conducted as part of the authors’ routine employment at BEHRI, Dhaka, Bangladesh.

## Acknowledgments

The authors thank the nurses, ophthalmic assistants, and optometrists of Bashundhara Eye Hospital and Research Institute (BEHRI) for their contributions to patient screening and data collection during the outreach eye camps.

## Notes

### Competing Interest Statement

The authors have declared no competing interest.

### Author Declarations

IRB of Bashundhara Eye Hospital and Research Institute waived ethical approval for this work

