## Supplemental table for "Ocular diagnostic patterns among patients attending community outreach eye camps in Bangladesh: a retrospective registry-based study"

**Supplementary table 1:** Frequency distribution of original registry diagnostic entries among patients attending community eye camps (N = 7,267).

| **Original registry entries** | **n (%)** |
| --- | --- |
| Presbyopia | 2093 (28.80) |
| Age related cataract | 637 (8.77) |
| Refractive error | 1344 (18.49) |
| Conjunctivitis | 149 (2.05) |
| Dacryocystitis | 57 (0.78) |
| Blepharitis | 23 (0.32) |
| Dry eye | 182 (2.50) |
| Migraine | 58 (0.80) |
| Foreign body | 7 (0.10) |
| Pterygium | 182 (2.50) |
| Cataract | 888 (12.22) |
| Red eye | 147 (2.02) |
| Episcleritis | 12 (0.17) |
| Posterior subcapsular cataract | 26 (0.36) |
| Choroidal neovascularization | 2 (0.03) |
| Burning | 9 (0.12) |
| Maculopathy | 2 (0.03) |
| Corneal opacity | 9 (0.12) |
| Watering | 100 (1.38) |
| Headache | 94 (1.29) |
| Viral keratitis | 3 (0.04) |
| Phthisis bulbi | 2 (0.03) |
| Itching | 33 (0.45) |
| Discharge | 3 (0.04) |
| Black spot | 2 (0.03) |
| Myopia | 108 (1.49) |
| Allergic conjunctivitis | 420 (5.78) |
| Floaters | 9 (0.12) |
| Blurring | 3 (0.04) |
| Astigmatism | 20 (0.28) |
| Entropion | 1 (0.01) |
| Pseudophakia | 122 (1.68) |
| Corneal ulcer | 6 (0.08) |
| Sac patency test | 36 (0.50) |
| Simple myopic astigmatism | 69 (0.95) |
| Compound myopic astigmatism | 30 (0.41) |
| Chalazion | 7 (0.10) |
| Systemic mastocytosis | 42 (0.58) |
| Stye | 1 (0.01) |
| Meibomian gland dysfunction | 7 (0.10) |
| Simple hypermetropia | 14 (0.19) |
| Madarosis | 1 (0.01) |
| Sinusitis | 7 (0.10) |
| Conjunctival lymphoma | 1 (0.01) |
| Pinguecula | 1 (0.01) |
| Trauma | 1 (0.01) |
| Glaucoma | 14 (0.19) |
| Ptosis | 3 (0.04) |
| Blind eye | 1 (0.01) |
| Squint | 7 (0.10) |
| Congenital nasolacrimal duct obstruction | 3 (0.04) |
| Bell’s palsy | 1 (0.01) |
| Ocular allergy | 53 (0.73) |
| Distance vision | 7 (0.10) |
| Subconjunctival hemorrhage | 1 (0.01) |
| Exotropia | 1 (0.01) |
| Irritation | 8 (0.11) |
| Eye ache | 10 (0.14) |
| Immature cataract | 32 (0.44) |
| Simple hypermetropic astigmatism | 5 (0.07) |
| Fungal | 2 (0.03) |
| Stone eye | 1 (0.01) |
| Photophobia | 3 (0.04) |
| Low vision | 26 (0.36) |
| Hypermetropic astigmatism | 4 (0.06) |
| Injury | 2 (0.03) |
| Corneal scar | 4 (0.06) |
| Vernal keratoconjunctivitis | 2 (0.03) |
| Hypermature cataract | 3 (0.04) |
| Optic atrophy | 1 (0.01) |
| Aphakia | 3 (0.04) |
| Retinal dystrophy | 1 (0.01) |
| Chronic uveitis | 2 (0.03) |
| Poor vision | 49 (0.67) |
| Emmetropia | 6 (0.08) |
| Hyperopia | 42 (0.58) |
| **Total** | **7267** |

**Supplementary Table 2:** Mapping of original registry diagnostic entries to the five analytic diagnostic categories used in this study.

| **Diagnostic category** | **n (%)** | **Original registry entries included** |
| --- | --- | --- |
| Cataract and lens disorders | 1711 (23.54) | Cataract, Age related cataract, Posterior subcapsular cataract, Immature cataract, Hypermature cataract, Pseudophakia, Aphakia |
| Refractive errors and presbyopia | 3729 (51.31) | Presbyopia, Refractive error, Myopia, Hyperopia, Simple myopic astigmatism, Compound myopic astigmatism, Astigmatism, Simple hypermetropia, Simple hypermetropic astigmatism, Hypermetropic astigmatism |
| Ocular surface inflammatory disorders | 624 (8.59) | Allergic conjunctivitis, Conjunctivitis, Ocular allergy, Vernal keratoconjunctivitis |
| Lacrimal system disorders | 196 (2.70) | Dacryocystitis, Watering, Sac patency test, Congenital nasolacrimal duct obstruction |
| Miscellaneous ocular disorders | 1007 (13.86) | Other conditions |
| **Total** | **7267** |  |
